# Barriers influencing the care of survivors of gender-based violence in the context of forced displacement in Kaya, Burkina Faso: a qualitative study

**DOI:** 10.1101/2024.04.01.24305078

**Authors:** Souleymane Bayoulou, Patrice Ngangue, Soutongnoma Safiata Kaboré, Boris Arnaud Kouomogne Nteungue, Yacouba Pafadnam, Danièle Sandra Yopa, Josiane Seu, Gbètogo Maxime Kiki

## Abstract

Gender-based violence (GBV) is a serious international health problem that challenges fundamental human rights. In addition to violating fundamental rights, it is an attack on the principles of gender equality. GBV is exacerbated in situations of conflict and forced displacement because of the vulnerability it engenders, particularly among women and children. Therefore, caring for victims of GBV in the context of a humanitarian crisis is special because of the specific nature of the context itself. This study aimed to explore the factors that negatively influence the care of survivors of GBV in the context of forced displacement. This was a descriptive and analytical-qualitative study. A total of 26 participants participated in the individual interviews. Most participants were healthcare workers (53,8%). The findings show that barriers to care are sociocultural (fear of stigmatization, ignorance of the benefits of seeking appropriate care and support, self-censorship among GBV survivors, fear or risk of reprisal, honor denied, fear of the aggressor punishment, shame and fear of being repudiated), institutional (lack of qualified human resources to care for GBV survivors, insufficient information on the availability of care services, geographical inaccessibility of care services, lack of confidentiality from service providers, inadequate health care and support systems, low availability of care services, poor quality of care) and financial. Bringing all care services together in one place, such as a one-stop center, is imperative to overcome these obstacles. In this way, it will be possible to improve accessibility to services and enhance the quality of care through effective coordination.

## INTRODUCTION

At the end of 2022, 108.4 million people worldwide were forcibly displaced from their home countries or towns, leading to a significant amount of 35.3 million refugees, 76% of whom were hosted in low- and middle-income countries [1]. Forced population movements exacerbate the risk of gender-based violence (GBV) [2].

GBV refers to any harmful action taken against an individual or group because of their gender identity [3]. It is a serious international health problem that questions fundamental human rights. In addition to violating fundamental rights, it is an attack on the principles of gender equality [4]. It is, therefore, a violation of human rights and affects not only the people who are its direct victims but also society as a whole [5].

GBV remains one of the world’s most widespread scourges, and its impact is far-reaching [6]. However, it is important to note that, as a general rule, the number of people who have reported acts of GBV, but not the number of people who have experienced it, is known [7]. In other words, the prevalence of GBV is extremely difficult to determine, as the concealment of many cases leads to underreporting. In contrast, it has been established that GBV is a widespread problem concerning human rights and public health [7].

Individuals can experience GBV, including men, boys and sexual and gender minorities [8]. It is a universal phenomenon and involves multisectoral management [6]. Nevertheless, women and girls are disproportionately affected [8]. Their status as women and girls in this crisis context increases their vulnerability and exposes them to these harmful practices [9]. Unfortunately, emergencies increase the risk of violence, exploitation and abuse [10]. Indeed, in the context of a crisis, due to a lack of resources, capacity and political will, women are more exposed to sexual and gender-based violence (SGBV) [5].

Burkina Faso (BF) has faced political, security and humanitarian crises since 2015. According to National Emergency and Rehabilitation Council (CONSUR) data, nearly two million people have been forcibly displaced to save their lives [2]. This situation is more marked in the Centre-Nord region, where thousands of people are forced to flee their homes [12]. According to the latest CONASUR statistics released on March 31, 2023, the Centre-Nord region has 493,945 internally displaced persons (IDPs), mostly women and children [13].

According to the GBV subcluster report, 2,393 cases of GBV were reported in 2022, compared with 1,197 cases in 2021[14]. At the same time, BF is witnessing a reduction in humanitarian space, with a profound malfunctioning of essential social services with access limitations for the most vulnerable categories of the population, including women, children, people with disabilities and older people [15]. All these situations are GBV multipliers and have a major impact on the sexual and reproductive health (SRH) of women and girls [16].

Humanitarian actors stress the need to provide urgent assistance to survivors of sexual and gender-based violence to ensure their protection and treatment and to avoid the discrimination they still face [17]. In parallel with this humanitarian crisis, cases of gender-based violence (GBV) have multiplied, with health, social, psychological and economic consequences [15]. Gender-based violence is a serious international health problem that challenges fundamental human rights. In addition to violating basic human rights, it undermines the principles of equality between men and women [4].

This study explored factors that negatively influence GBV survivors’ use of care among IDPs in the Centre-Nord region of Burkina Faso.

## METHODS

### Study design

We conducted a qualitative, holistic, and analytical case study to analyze the care use of IDPs.

### Study population

The study population is composed of three (03) target groups: (i) GBV survivors, (ii) professionals involved in the management of GBV survivors (health workers, psychologists, case managers and criminal investigation officers), and (iii) coordinators of management services.

### Inclusion criteria GBV survivors

To be included in the study, participants had to be IDP living in Kaya, survivor of GBV who received care and were present in Kaya at the time of the study.

### Sampling

We conducted stratified random selection to ensure that diversity could be found within the sample. Our sample was distributed as follows:

- 11 survivors of GBV: For this target group, we considered all GBV survivors who had received care and had given their free and informed consent to be interviewed.

Twenty-six people were involved in the psychological, medical and legal care of GBV survivors. They come from the care services in the zones to welcome displaced persons. They include health workers, psychologists, social workers, case managers and Judicial Police Officers (OPJ).

- two coordinators of care services.

### Data collection methods, techniques and tools

Two techniques were used to collect the data: individual interviews and document review. The semi-structured interview guide for each group of participants was deployed on smartphones using a Kobotoolbox. Individual interviews were conducted in the services concerned. All open-ended questions were systematically audio recorded using the KoboCollect tool, with the interviewees’ consent. Each interview lasted an hour on average. The interview guides were tested under real-life conditions to verify their validity and reliability and assess their relevance and average completion time before validation and corrections were made. Three pairs of trained and experienced interviewers conducted the interviews. The training was practical in enabling participants to familiarize themselves with the KoboCollect tool and, above all, to gain a shared understanding of the questionnaire in the local language spoken by internally displaced persons (IDPs).

### Data analysis

Sociodemographic characteristics from individual interviews with care providers were coded in Microsoft Excel and analyzed using the Statistical Package for the Social Sciences software SPSS version 25, SPSS Inc. Armonk, NY: IBM Corp.

We opted for the qualitative thematic analysis proposed by Braun and Clarke (2006), an inductive approach to identify and analyze themes emerging from the participants’ entire discourse [18]. Unlike other approaches, it does not impose predetermined categories based on a theoretical framework [19]. Consequently, we did not precategorize the responses [20]. To carry out the analysis, we followed the six steps described by Braun and Clarke as follows (1) free coding ; (2) classification of meaning units ; (3) creation of categories and subcategories ; (4) theme development ; (5) theme mapping ; (6) presentation of results.

### Ethical considerations

The Burkina Faso Health Research Ethics Committee approved the study protocol under number 2022-01-012 on January 17, 2022. All participants were informed of the study’s objectives and the possibility of withdrawing from the study at any time without prejudice. The purpose of the audio recording was also clearly explained to them in the local language. Additionally, each participant provided informed consent before the interview with the assurance that their answers were anonymized for confidentiality purposes. Moreover, the tools used did not allow direct identification of study participants.

## RESULTS

A total of 26 participants participated in the individual interviews. Most participants were healthcare workers (53,8%), as shown in Table 1 below with more details.

**Table 1.**
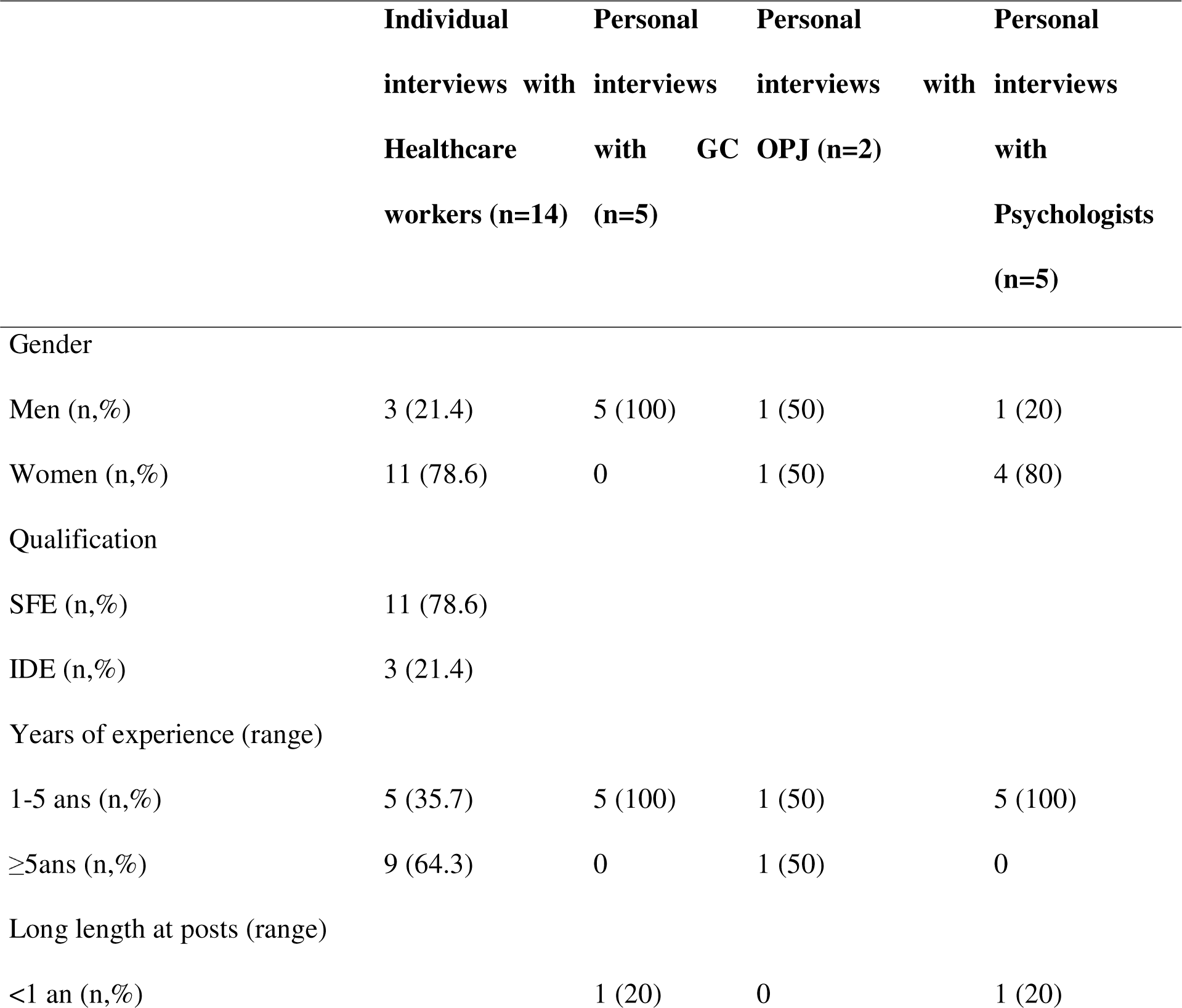
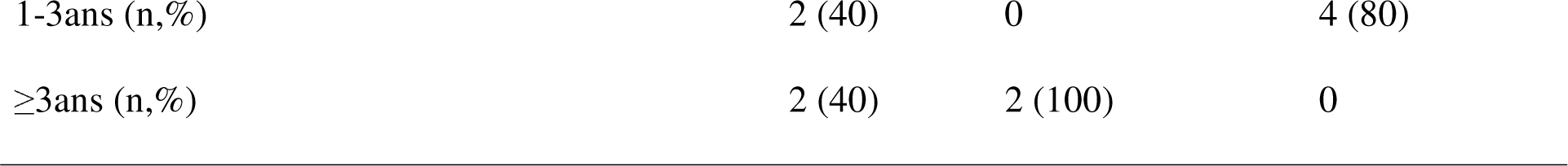
Sociodemographic characteristics of GBV care providers in Kaya, Burkina-Faso (n=26)

Victims need holistic and appropriate care to rebuild their lives. However, there are often barriers that prevent survivors from accessing intake services. The results of the study highlighted three types of barriers: (i) sociocultural barriers, (ii) institutional barriers and (iii) financial barriers.

### Sociocultural barriers

The results of the study show that the main sociocultural barriers are as follows:

#### Fear of stigmatization

Individual interviews with care providers also showed that the fear of being stigmatized is a real barrier to care: *“…stigmatization, rejection, shame, the way society looks at them can be reasons for a survivor not to want to report GBV” (Individual interview with psychologist 2)*. Indeed, some survivors are afraid of being stigmatized by reporting their GBV situation to care services. They are so scared of not being understood and of being marginalized.

We also note that more than most care providers (16/26) believe that women are responsible for the violence they experience due to their behavior. Additionally, almost half (12/26) believe that the victims of GBV do not consider injuries inflicted by their husbands to be GBV. In other words, survivors often suffer double violence. They are sentenced without trial. This is a clear example of how to reinforce victims’ conviction that there is no point in seeking help from these services. In many cases, acts of GBV are perceived as part of the socialization process and are therefore encouraged by society, so much so that laws to combat GBV are seen as a breach of the social fabric. Even those who are supposed to take action to eliminate GBV are involved in spreading preconceived and misleading ideas.

#### Ignoring the benefits of seeking appropriate care and support

Some survivors are unaware of their rights: “*Others do not know that after a rape, you have to go to a health center” (Individual interview, health agent 12)*. They suffer from GBV without knowing that their rights are being violated: “*Some people are victims of GBV, but they do not know that it is GBV. Take the example of a woman who lives with her husband and who forces her to have sex at any time, even though she is not consenting at the time. She may not know this is GBV” (Individual interview, case manager 3)*.

Under these conditions, they take no initiative to escape the cycle of violence, let alone seek help for appropriate care. The study also revealed that the vast majority of GBV survivors have no formal education. This state of affairs could exacerbate their vulnerability and contribute to keeping them in the cycle of violence.

Similarly, half of the stakeholders (13/26) believe that unequal relations between men and women are not risk factors for gender-based violence.

#### Self-censorship among GBV survivors

Some survivors believe that they are responsible for their misfortune: “*One obstacle is that GBV survivors tell themselves that there is no longer any possibility of someone helping them to overcome what happened to them because the past act is now part of their lives. It is a form of resignation” (Individual interview with Psychologist 1)*. This situation plunges them into absolute silence because they are convinced there is no way to help them in the face of the evidence they are going through. For them, the past is now part of their lives, and they will have to live with it in their consciences. This kind of resignation is a real obstacle to taking charge of their lives.

#### Fear or risk of reprisal

Very often, the aggressor lives in the victim’s family environment. This may be an intimate partner, a care provider, a family member, or an acquaintance. Given the perpetrator’s social status, this proximity can hinder reporting. She may also be pressured, threatened or killed by her attacker to avoid humiliation. In such cases, the survivor prefers to keep herself secret and suffer martyrdom: “*A survivor may not report her GBV because if the person who assaulted her is someone close to her, an acquaintance, a relative, she will be afraid that by reporting this violence to the authorities, she may be marginalized” (Individual Interview Case Manager 4)*.

#### Honor denied

According to healthcare providers, survivors are often afraid that their honor will be trampled after they report GBV: “*Then, they think that their honor has been trampled” (individual interview with healthcare provider 11)*. As a result, they prefer not to report GBV, which is a barrier to care.

#### Fear of the aggressor’s punishment

In many situations, women survivors of GBV resign themselves to enduring the violence because they want to protect their tormentor: “*A woman who her partner beats, if she reports it to the police, they will go straight to extreme punishment, even imprisoning the husband” (Individual interview, case manager 1)*. Indeed, in situations of intimate partner violence, survivors, for fear of seeing their spouse in prison, simply decide not to talk about it and to suffer in silence.

#### Shame

In some cases, such as incest, victims feel obliged to remain quiet to avoid shame: “*The main reason for me is to feel this sense of shame after having been subjected to such a situation” (Psychologist 3, individual interview)*. These kinds of abominations are not always disclosed to protect the family from marginalization; for example, “*I take the case of incest; someone who has slept with his daughter or the brother who has slept with his sister or with a family member. These kinds of cases can never be disclosed because the family will never accept them, and they will stay between them. Unless there’s a case of illness or pregnancy, and now, they often use our service a little clandestinely to help them find a solution” (Individual interview, case manager 5)*.

#### Fear of being repudiated

In cases of intimate partner violence, many victims resign themselves to speaking out to avoid repudiation: “*Women who live in couples, even if they are mistreated, do not like to complain because the husband may now decide to expel her, so they put up with it.” Individual interviews with case managers 5*. “*Some women, because of their children, are afraid of being repudiated by their husbands” (Individual interview with Judicial Police Officer 1)*. Unfortunately, tragedies also occur *under these conditions*.

### Institutional barriers

The results of the study identified eight (n=08) types of institutional barriers to the care of GBV survivors:

#### Lack of qualified human resources to care for GBV survivors

Care providers’ technical and operational capacities are weak: “*The difficulty of finding specialized human resources to ensure adequate care is genuine” (MCD individual interview)*. The study revealed that some care providers are unfamiliar with the conditions of care, the chronology of treatment, and the deadline for administering postexposure prophylaxis (PEP) and emergency contraception in the event of rape. As a result, some health workers are reluctant to take on cases of rape because they do not know exactly what to do: “*Some health workers are afraid to take on cases of GBV because they are not trained” (Interview with an individual health worker)*.

For example, the vast majority of health agents interviewed (10/14) believe that it is mandatory to examine the victims of sexual violence before administering treatment. Of the 14 health workers interviewed, not one was able to list in chronological order the priorities for treatment in cases of rape. Slightly more than one-third (5/14) of health workers thought knowing the victim’s serological status before administering postexposure prophylaxis (PEP) was essential. Nearly half (6/14) of health workers are unaware that the maximum time limit for administering PEP is 72 hours.

More than half (10/14) of the health workers interviewed were unaware that the maximum time limit for administering emergency contraception (EC) was five (5) days. In addition, almost half (6/14) of the mothers did not know other nonsurgical methods to avoid pregnancy after sexual violence when emergency contraceptive pills were unavailable. Most health workers (9/14) believe that the primary purpose of physical examination is to determine with certainty whether rape has occurred or to determine whether prophylaxis for sexually transmitted infections (STIs) is necessary rather than to assess and document injuries.

Regarding capacity building, one-third of care providers have never been trained on a topic related to GBV. Most of the players interviewed were unaware of the guidelines for the management of GBV survivors. According to the majority (15/26), the minimum response in the safe care of the survivor is to stop the violence immediately rather than to guarantee the victim’s safety and security. In addition, almost half (12/26) believe that suicide-related issues should not be discussed with GBV survivors. In addition, more than half (15/26) feel the victim must tell her story before being treated. In addition, one-third of those involved (9/26) believed that the victim’s informed consent should be sought during the treatment. However, it should be noted that all care providers are unanimous in saying that survivors must receive fair and equitable treatment, regardless of their ethnic origin, religion, nationality or sexual orientation.

#### Insufficient information on the availability of care services

There is a lack of awareness of existing care services due to a lack of communication: “*The fact that there are not many broadcasts or talks on the subject means that people are not informed” (Individual interview, case manager 3)*.

Indeed, there are very few radio broadcasts and other awareness-raising activities organized by care services to orient and enlighten the population: “*If they are not informed that these structures exist, they will stay at home with their problem” (Individual interview, case manager 4)*. Thus, the lack of information is often the cause of the ignorance of many people, who do not even know that they are victims of human rights violations. The study also revealed that even within the same department, not all players are at the same level of information. For example, some actors (2/26) had no information on the existing referral circuit. This lack of information can lead to delays in referral and, consequently, negatively impact the quality of care. Moreover, most survivors felt they had not received information on available services and options. One-third of the survivors thought they had not received a referral, even though the service was unavailable locally.

#### Geographical inaccessibility of care services

The study showed that even when the service is available, survivors are not always aware of its geographical location: “*It also depends on the location of the service. If the service is not well located, the person risks exposing himself to the view of his community because some victims may think that everyone sees them when they enter the service” (Individual interview, case manager 2)*. It also emerges that there is a problem of geographical accessibility because the areas where internally displaced persons (IDPs) congregate are in the outlying districts, whereas the services are more centrally located: “*There are geographical obstacles; this means that the person is in a corner that is far from the health facility; he often has to cross low-lying areas where there is insecurity” (Individual interview with health agent 10)*. Indeed, more than one-third of the survivors interviewed (4/11) indicated that the services are difficult to find. On the other hand, all the survivors felt that the opening hours of the services enabled them to access them without difficulty or constraints.

#### Lack of confidentiality from service providers

Care providers have indicated that one of the barriers to care is the lack of ethics and deontology on the part of certain care providers: “*There is also the very credibility of the service; the agents who work in the service must know how to demonstrate ethics and deontology, respect for the victim’s dignity” (Individual Interview Case Manager 2)*. “*In my opinion, there’s also the judgment passed on the care services, which is considered unsafe for her and the lack of trust toward the care providers (Individual Interview Psychologist 1)*. Unfortunately, some staff may go so far as to betray professional secrecy out of sheer thoughtlessness. Prejudices about those involved in the care process are also seen as a further obstacle to requesting services: “*I think many people are afraid to approach us because they’re simply prejudiced against women who wear the uniform” (Individual Interview Police Officer 1)*.

#### Inadequate health care and support systems

Direct observation has shown that almost all health care services do not have a suitable setting for receiving GBV survivors with complete confidentiality: “*We lack a suitable setting for receiving victims of gender-based violence” (individual interview with health worker 5)*. The infrastructure is ill-suited to providing better care conditions for survivors of GBV. One-third of the patients (10/26) did not even have room to ensure the confidentiality and dignity of the survivors throughout the treatment.

#### Low availability of care services

The low availability of care services is linked to the quiet operational presence of care providers and the closure of health centers due to insecurity: “… *it is the very availability of the service, if the agents who are authorized to look after the person are not available, it is very complicated!” (Individual interview, case manager 4)*. Moreover, the geographical coverage of care services is disparate due to the reduction in humanitarian space, which also impacts access to care services. ***Poor quality of care***

The individual interviews revealed that the poor behavior of certain staff in care services could be an obstacle to care: “*It is the obstacle linked to the quality of care. For example, if care does not go well, you have to understand that this can encourage other people not to come forward” (Individual interview with health worker 5)*. These attitudes can affect the quality of care and even create secondary trauma for the victim. Two-thirds of the survivors felt that the services were not welcoming. Interview with a health worker: “Poor reception by health workers can lead a GBV survivor not to use our health services” *(Interview with a health worker 9)*.

#### Lack of cross-sector collaboration

Collaboration between stakeholders is not always effective, even though it is imperative to respond adequately to the survivor’s needs, as well as to mitigate the risks of GBV in other sectors, because the issue is cross-cutting: “*If the agent who provides the care does not collaborate with other services to find appropriate solutions to the needs of the victim of gender-based violence, it would be difficult” (Interview health agent 3)*.

### Financial barriers

At the health center level, an analysis of the narratives of health workers who care for GBV victims revealed that some of them believe that GBV victims do not always seek treatment because they think that the costs associated with their treatment and follow-up would be high and that they could therefore not afford it: “*Many say that they cannot afford treatment and the procedures can be long to receive care. Maybe they do not know that if they came, we could help them” (Individual interview with health agent 14)*.

Thus, a lack of financial means can be a barrier to treatment. Moreover, some victims are tempted to believe that cost has an additional impact on increasing the length of procedures and associated treatments. However, very few players (2/26) mentioned this possibility. Indeed, apart from financial support from United Nations agencies and international nongovernmental organizations (NGOs), the treatment of cases of gender-based violence (GBV), particularly cases of sexual violence requiring surgical repair, is not always free of charge. A medical certificate costs an average of five thousand (5,000) CFA francs.

## DISCUSSION

This study has enabled us to gain a better understanding of the factors hindering access for survivors of GBV in the context of forced displacement. Indeed, an analysis of the management mechanism reveals that the obstacles are mainly at three levels. First, there is the victim’s sociocultural environment and stereotypes. The sociocultural burden often forces victims to resign and not seek help [21]. Moreover, some obstacles are intrinsically linked to care services. The gap is more significant in a context marked by a shortage of qualified human resources, low expertise among care providers, poor availability and quality of services, and poor quack of credibility on the part of care services [7]. Finally, there is a cost of care, which is not always affordable for victims. The results closely reflect global data and findings from other humanitarian contexts [22]. In terms of sociocultural barriers, studies have shown that the reasons for not reporting gender-based violence (GBV) and refusing to seek help are generally linked to fear of social rejection, shame, ignorance, fear and the presence of the perpetrator in the victim’s living environment [23]. Stigmatization plays a crucial role in women’s decision to seek help from GBV services [22]. Survivors are often ashamed and fearful of being rejected by society, especially when the perpetrator lives in their immediate environment. These attitudes prevent them from reporting GBV or seeking help. At the 57th session of the United Nations Economic and Social Council held in 2013, the program coordinator for victims of violence at Argentina’s Ministry of Justice and Human Rights explained that despite the application of a very comprehensive law adopted in her country to eliminate violence against women, the latter still experienced fears of “reprisals” in the event of a complaint [24]. Care providers who are supposed to protect victims are also often alienated by stereotypes and preconceived ideas [25]. For example, more than a majority of the care providers interviewed believe that women are responsible for the violence they suffer as a result of their behavior, and more than a third believe that men’s insults to their wives are not GBV. Under these conditions, it seems clear that many of those involved in the care of survivors will not be able to help them put their problems into words, let alone avoid feeling guilty or condemning themselves. Unfortunately, victims construct their experiences through laws and social policies referred to by various institutions, such as family or community organizations [26]. In terms of institutional barriers, we note that healthcare providers generally have a low level of expertise in caring for victims of sexual and other forms of gender-based violence (GBV) [27]. In humanitarian contexts, survivors of GBV must receive prompt care. Although support services may be available, maximizing their benefits remains challenging [22]. The study showed that there is a real shortage of qualified human resources at all levels of care. Very few of those involved in care are trained.

Moreover, there is a high turnover rate due to the security situation. As a result, many trained professionals, particularly health workers, ask to be posted to other regions considered safer. This situation negatively impacts the quality of care, even if referrals are well made from the outset. As a result, victims are often confronted with poor reception and inadequate care. The study revealed that most health workers are unaware of the treatment protocol in the event of rape. Some health workers do not know the priorities for treatment, care conditions, guidelines, or even the referral mechanism.

Similarly, psychologists are unaware of the different needs to be assessed in a survivor, let alone the dimensions of mental health and psychological support response [28]. In addition to needs assessment, the study revealed that GBV case managers do not master the steps involved in case management. Survivors also pointed to a lack of professionalism by those involved. Most survivors did not appreciate how they received care services. Some even questioned the lack of respect for confidentiality, one of the guiding principles of the services provided. The study also revealed that the closure of health facilities due to growing insecurity hinders access to care and makes it difficult to gather feedback information to guide decision-making [27].

Health services are the main gateway for survivors to receive care services. This healthcare system failure can result in less access to sexual and reproductive health (SRH) services, social services and justice [4]. Even the services available are not all easily accessible (physical accessibility). More than one-third of the survivors interviewed mentioned this difficulty. We also note that, even when the service exists, the availability of confidential spaces and qualified male and female staff is only sometimes a reality. The poor behavior of certain care providers was also identified as an obstacle to care. Inadequate communication about the presence of care services could also explain survivors’ difficulties accessing such services. None of the care providers mentioned their responsibility to respect the principle of confidentiality or their obligation to communicate that services are free. All these factors constitute obstacles to accessing services and contribute to lengthening the time it takes to receive care.

A lack of financial resources is another barrier to treatment. Indeed, treatment is not always free of charge. In addition, this financial barrier, which delays treatment, can render it ineffective in certain situations, notably in cases of rape, where prophylaxis for acquired immunodeficiency virus (HIV) must imperatively be carried out within 72 hours of the act of rape [29]. A small number of respondents recognized financial barriers because of the availability of subsidized care by the government and international organizations in the studied healthcare facilities, allowing the reduction of finances as a strong barrier for survivor care. Moreover, the cost of services, if not free, must be accessible to all survivors of GBV [30].

This study has several limitations. Reporting bias may be linked to the subject’s sensitivity, leading some actors to underreport by refraining from mentioning certain sensitive or hurtful aspects [31]. However, mobilizing all stakeholders in the fight against GBV is difficult without conclusive data and scientific evidence [32]. Hence, the scientific interest in this topic is high.

## CONCLUSION

We have demonstrated that obstacles to adequate, holistic care for GBV survivors are linked to organizational dysfunctions and sociocultural constraints. However, without effective coordination, it will be easier to respond holistically to the needs of GBV survivors. This is why we need to raise awareness of vulnerable groups of GBV victims and develop integrated care centers. The aim is to make the full range of management activities available in one location with funds. This has the advantage of facilitating access to services and enhancing the quality of care thanks to a well-coordinated response.

## Data Availability

All data produced in the present work are contained in the manuscript

